# Factors Contributing to Healthcare Professional Burnout During the COVID-19 Pandemic: A Rapid Turnaround Global Survey

**DOI:** 10.1101/2020.05.17.20101915

**Authors:** Luca A. Morgantini, Ushasi Naha, Heng Wang, Simone Francavilla, Ömer Acar, Jose M. Flores, Simone Crivellaro, Daniel Moreira, Michael Abern, Martin Eklund, Hari T. Vigneswaran, Stevan M. Weine

## Abstract

**Background:** Healthcare professionals (HCPs) on the front lines against COVID-19 may face increased workload, and stress. Understanding HCPs’ risk for burnout is critical to supporting HCPs and maintaining the quality of healthcare during the pandemic.

**Methods:** To assess exposure, perceptions, workload, and possible burnout of HCPs during the COVID-19 pandemic we conducted a cross-sectional survey. The main outcomes and measures were HCPs’ self-assessment of burnout and other experiences and attitudes associated with working during the COVID-19 pandemic.

**Findings:** A total of 2,707 HCPs from 60 countries participated in this study. Fifty-one percent of HCPs reported burnout. Burnout was associated with work impacting household activities (RR=1·57, 95% CI=1·39-1·78, *P*<0·001), feeling pushed beyond training (RR=1·32, 95% CI=1·20-1·47, *P*<0·001), exposure to COVID-19 patients (RR=1·18, 95% CI=1·05-1·32, *P*=0·005), making life prioritizing decisions (RR=1·16, 95% CI=1·02-1·31, *P*=0·03). Adequate personal protective equipment (PPE) was protective against burnout (RR=0·88, 95% CI=0·79-0·97, *P*=0·01). Burnout was higher in high-income countries (HICs) compared to low- and middle-income countries (LMICs) (RR=1·18; 95% CI=1·02-1·36, *P*=0·018).

**Interpretation:** Burnout is prevalent at higher than previously reported rates among HCPs working during the COVID-19 pandemic and is related to high workload, job stress, and time pressure, and limited organizational support. Current and future burnout among HCPs could be mitigated by actions from healthcare institutions and other governmental and non-governmental stakeholders aimed at potentially modifiable factors, including providing additional training, organizational support, support for family, PPE, and mental health resources.

**Funding:** N/A

## Research in context

### Evidence before this study

The authors conducted a search on the PubMed search engine from 3/1/2020 to 3/10/2020 with the terms “COVID-19” (replaced also with the related terms “SARS-CoV-2”, “coronavirus”, and “pandemic”) AND “burnout” AND “healthcare.” The results of the search, not limited to the English language, were reviewed by the authors within the same timeframe. All evidence published in peer-reviewed journals was reviewed by the authors.

### Added value of this study

Our study is the first worldwide survey of healthcare professionals during the COVID-19 pandemic and demonstrates how burnout is prevalent at higher than previously reported rates. Burnout was found to be related to several modifiable factors, including the availability of additional training, organizational support, family-related support, personal protective equipment, and mental health resources. Reported burnout was higher in high-income countries compared to low- and middle-income countries

### Implications of all the available evidence

Our findings offer insight into the unique impact of this pandemic on healthcare professionals across the globe. Policymakers and other governmental and non-governmental stakeholders will be able to better understand how to mitigate current and future burnout among healthcare workers that are on the front lines against COVID-19.

## Introduction

More than 200 countries worldwide are impacted by the spread of the novel coronavirus (COVID-19). Their healthcare systems are maximizing efforts in order to deploy resources to mitigate spread and reduce morbidity and mortality from COVID-19.

Large numbers of healthcare professionals (HCPs) on the frontlines face high adversity, workloads, and stress, making them vulnerable to burnout.^1,2^ Burnout, defined as emotional exhaustion, depersonalization, and low personal achievement, is known to detract from optimal working capacities, and has been previously shown to be prevalent among HCPs across the globe with a similar rate in high and low income countries^3^. Burnout has been found to be driven by high job stress, high time pressure and workload, and poor organizational support.^3^

The objective of this study was to understand the impact of COVID-19 on HCPs working during the pandemic, from a global perspective. We aimed to describe contributing factors associated with HCPs burnout.

## Methods

### Human Subjects Research

The University of Illinois at Chicago (UIC) Institutional Review Board determined on April 1^st^, 2020 that this study, with the assigned protocol number 2020-0388, met the criteria for exemption as defined in the U.S. department of Health and Human Services Regulations for the Protection of Human Subjects [45 CFR 46. 104(d)]. Before initiating the survey, respondents were informed that their responses would be shared with the scientific community. Survey responses were recorded and stored without participant identifiers using the REDCap electronic data capture software hosted by UIC servers. exhaustion, depersonalization, and low personal achievement, is known to detract from optimal

### Sample Population

Inclusion criteria was restricted to membership in COVID-19-specific social media groups restricted to HCPs. Platforms including Facebook, WhatsApp, and Twitter, as well as e-mail, were used for global recruitment and dissemination from April 6 to April 16, 2020. The survey was translated into 18 languages by professional translators.

### Outcomes and Measures

The survey contained 40 questions covering three major domains of HCPs experience (exposure, perception, and workload) that were validated by experts in infectious diseases, public health, occupational medicine, psychology, and clinical psychiatry. Elements of these domains were previously proposed as contributing toward HCP anxiety during the COVID-19 pandemic.^4^ The main outcome, HCPs-perceived burnout, was assessed by a single item on a 7-point Likert scale (1: strongly disagree to 7: strongly agree) as has been done in prior research, using the statement, “I am burned out from my work.”^5^ The questionnaire was developed with a pilot group of 10 HCPs and 40 questions were included based on expert opinion (Supplement 1) and then translated into 18 languages by professional translators. The country of the respondents was categorized as high-income or low- and middle-income as defined by the World Bank classification system.^6^ COVID-19 deaths and cases per 1 million population were obtained from a widely used web-based dashboard.^7^

### Statistical Analyses

A descriptive assessment was performed for each variable surveyed for all data, country by country and according to the income level (high vs. low-middle). Covariates collected as ordinal 5 scores variables were transformed into binary (Table 2, supplementary materials). For burnout, ≥5 were considered burned out. Quasi-Poisson regression analysis was performed using the binary burnout outcome.^8^ Relative risk (RR) was reported with nominal 95% confidence intervals and two-sided P-values. Participants who responded completely to the variables of interest were included in regression analyses.

## Results

A total of 2,707 valid responses were received from HCPs in 60 countries. Figure 1 demonstrates the study period in context of the COVID-19 pandemic (Figure 1; Table 3, supplementary materials).^6, 7^

**Figure 1:**
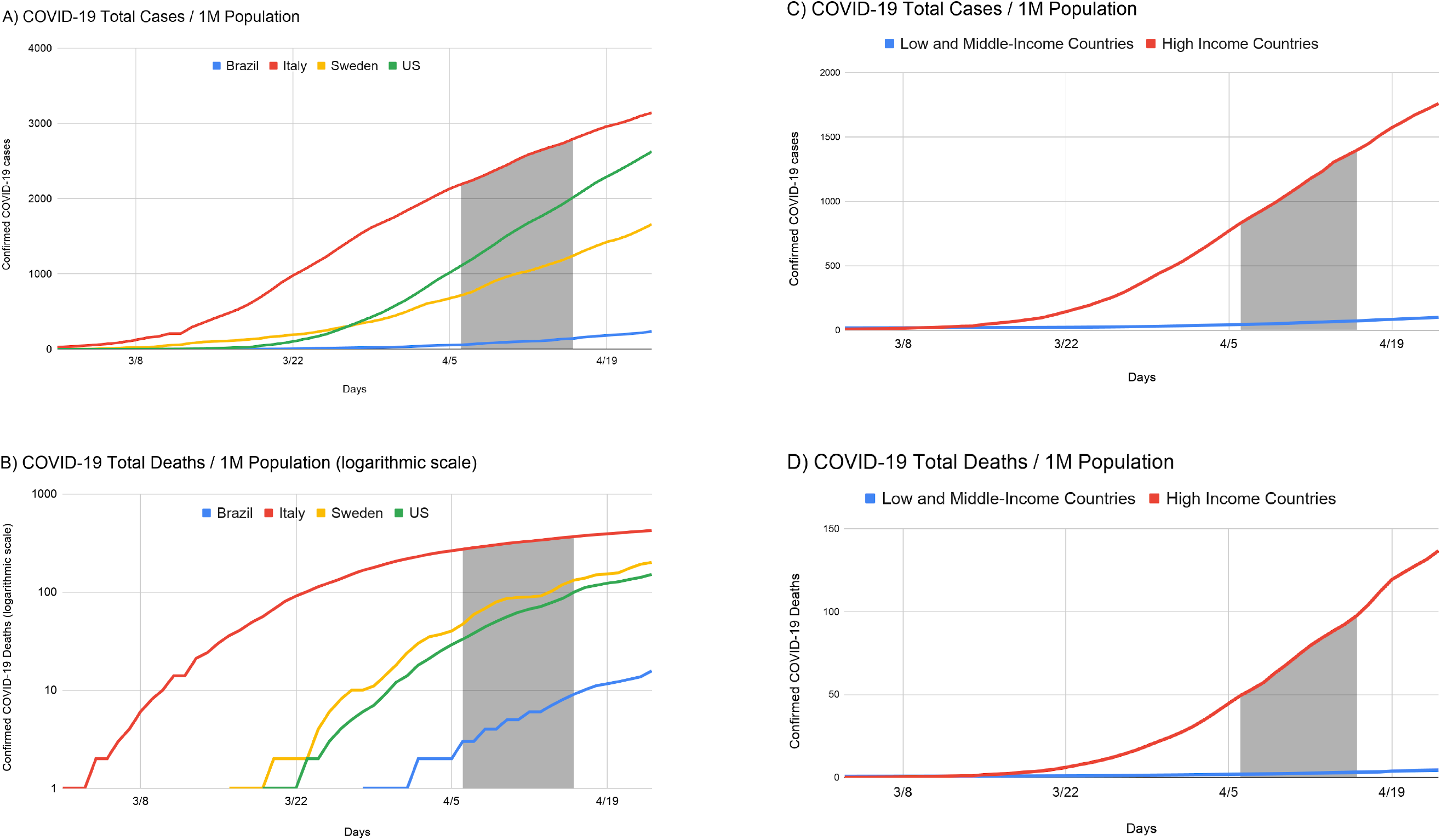
Total confirmed COVID-19 cases (A) and total confirmed COVID-19 deaths (B) per 1 million (M) population for the 4 countries with the highest response rates and for HICs (C) and LMICs (D).

Table 1 summarizes participant characteristics and responses (additional responses in Table 4, supplementary materials). Half (51·4%) of the respondents coming from 33 countries reported burnout because of their work during the COVID-19 pandemic.

**Table 1:** Healthcare professionals’ responses about perceptions, exposure, and workload during the COVID-19 pandemic. (PPE) Personal protective equipment; (QoL) Quality of life; (NP) Nurse practitioner; (PA) Physician assistant; (CRNA) Certified registered nurse anesthetist

|  |  |  |
| --- | --- | --- |
| **Country** |  |  |
|  | **Brazil** | 186 (6.9%) |
|  | **Italy** | 598 (22.1%) |
|  | **USA** | 833 (30.8%) |
|  | **Sweden** | 149 (5.5%) |
|  | **Other** | 941 (34.8%) |
| **Occupation category** |  |  |
|  | **Physician (Residents, Fellows)** | 719 (26.6%) |
|  | **Nurse (NP, PA, CRNA)** | 855 (31.6%) |
|  | **Other** | 1133 (41.9%) |
| **Exposed to a patient with COVID-19** |  |  |
|  | **No** | 644 (33.9%) |
|  | **Yes** | 1255 (66.1%) |
| **Symptoms suggestive of COVID-19** |  |  |
|  | **No** | 1526 (80.2%) |
|  | **Yes** | 377 (19.8%) |
| **Tested for COVID-19** |  |  |
|  | **No** | 1630 (85.7%) |
|  | **Yes** | 271 (14.3%) |
| **Positive test for COVID-19** |  |  |
|  | **No** | 221 (83.1%) |
|  | **Yes** | 45 (16.9%) |
| **Current perception of COVID-19** |  |  |
|  | **Benign disease** | 16 (0.9%) |
|  | **Mild disease** | 50 (2.9%) |
|  | **Moderate disease** | 534 (30.8%) |
|  | **Severe disease** | 1134 (65.4%) |
| **Adequate PPE was provided** |  |  |
|  | **No** | 778 (45.2%) |
|  | **Yes** | 945 (54.8%) |
| **Was mental health support available** |  |  |
|  | **No** | 902 (52.2%) |
|  | **Yes** | 825 (47.8%) |
| **Received COVID-19 specific training** |  |  |
|  | **No** | 921 (53.1%) |
|  | **Yes** | 815 (46.9%) |
| **Made life prioritizing decision** |  |  |
|  | **No** | 1470 (85.6%) |
|  | **Yes** | 248 (14.4%) |
| **Felt pushed beyond training** |  |  |
|  | **No** | 1174 (68.1%) |
|  | **Yes** | 550 (31.9%) |
| **Work impacting household activities because of COVID-19** |  |  |
|  | **No** | 500 (30.5%) |
|  | **Yes** | 1139 (69.5%) |
| **Work impacting QoL because of COVID-19** |  |  |
|  | **No** | 538 (32.8%) |
|  | **Yes** | 1100 (67.2%) |
| **I am burned out from my work (Likert 1-7)** |  |  |
|  | **Strongly disagree** | 146 (8.9%) |
|  | **Disagree** | 255 (15.6%) |
|  | **Somewhat disagree** | 114 (7.0%) |
|  | **Neither agree nor disagree** | 281 (17.2%) |
|  | **Somewhat agree** | 406 (24.8%) |
|  | **Agree** | 249 (15.2%) |
|  | **Strongly agree** | 187 (11.4%) |
| **I am burned out from my work (Binary)** |  |  |
|  | **No** | 796 (48.6%) |
|  | **Yes** | 842 (51.4%) |

Across all countries (Figure 2), reported burnout was associated with work impacting household activities (RR=1·57, 95% CI=1·39-1·78, *P*<0·001), feeling pushed beyond training (RR=1·32, 95% CI=1·20-1·47, *P*<0·001), exposure to COVID-19 patients (RR=1·18, 95% CI=1·05-1·32, *P*=0·005), and making life prioritizing decisions due to supply shortages (RR=1·16, 95% CI=1·02-1. ·1, *P*=0·03). Adequate personal protective equipment (PPE) was protective against reported burnout (RR=0·88, 95% CI=0·79-0·97, *P*=0·01). The likelihood of reported burnout was significantly higher in nurses compared to physicians (OR=1·47, 95% CI=1·12-1·92, *P*=0·006).

**Figure 2:**
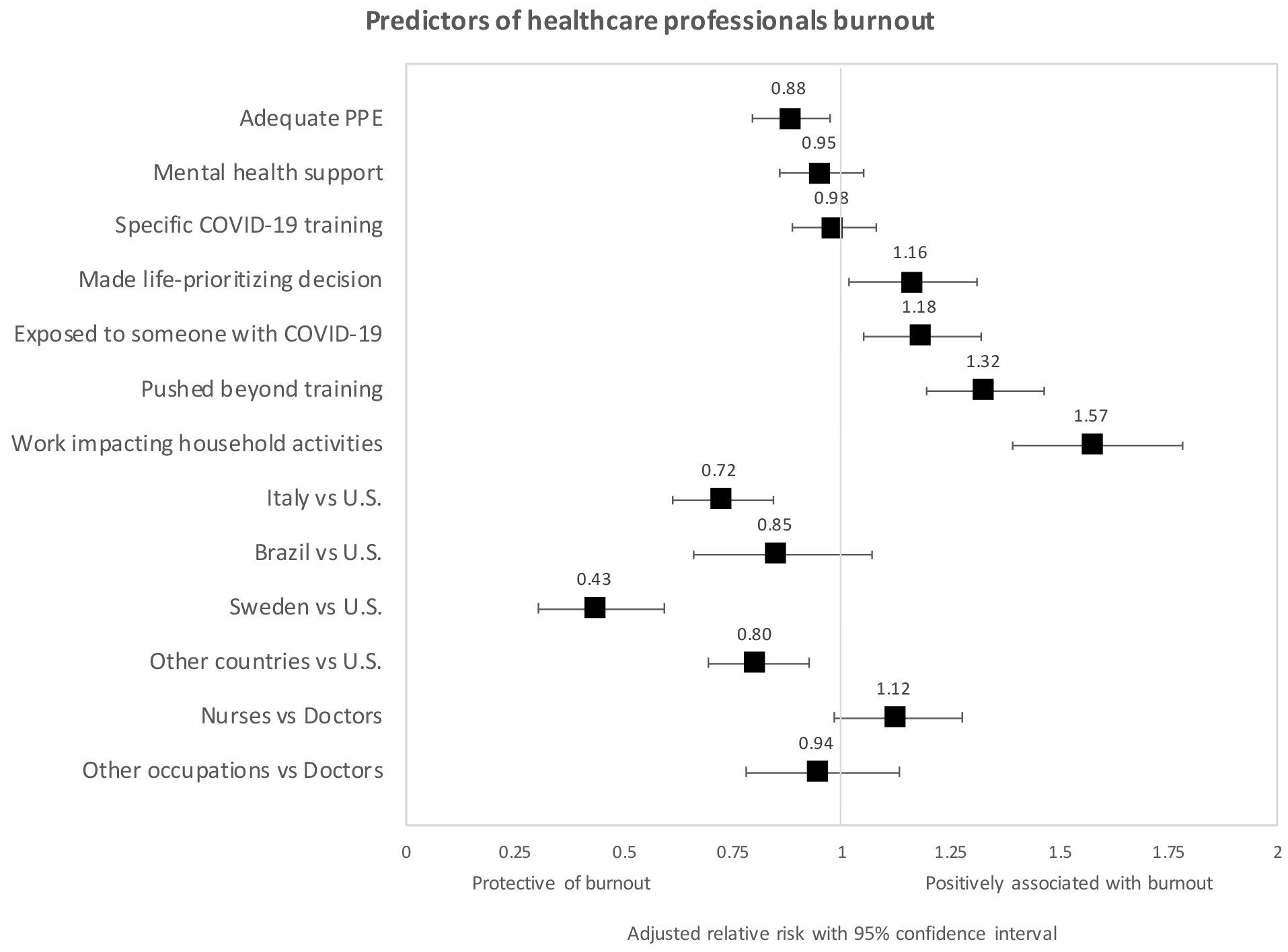
Forest plots shows adjusted relative risk (RR) for the multivariable regression analysis of burnout. (PPE) Personal protective equipment; (ICU) Intensive care unit; (ER) Emergency room; (ID) Infectious diseases.

Country-level analysis revealed lower reported burnout in Italy (RR=0·72, 95% CI=0·61-0·84, *P*>0·001) and Sweden (RR=0·43, 95% CI=0·30-0·59, *P*<0·001) compared to the United States (U.S.). Additionally, reported burnout was higher in HICs (High Income Countries) compared to LMICs (Low-to-Middle-Income Countries) (RR=1·18; 95% CI=1·02-1·36, *P*=0·018).

Predictors of burnout differed between LMICs and HICs (Figure 2, supplementary materials). In LMICs, reported burnout was associated with work impacting household activities (RR=2·31, 95% CI=1·61-3·43, *P*<0·001) and adequate PPE (RR=0·68, 95% CI=0·52-0·90, *P*=0·007). In HICs, reported burnout was associated with feeling pushed beyond training (RR=1·41, 95% CI=1·06-1·88, *P*=0·02), difficulty obtaining COVID-19 testing (RR=1·43, 95% CI=1·04-1·94, *P*=0·03), work impacting quality of life (RR=1·67, 95% CI=1·12-2·59, *P*=0·02), work impacting household activities (RR=1·75, 95% CI=1·16-2·75, *P*=0·01), and mental health support (RR=0·72, 95% CI=0·54-0·96, *P*=0·03).

## Discussion

Among respondents, half of HCPs from 33 countries reported burnout. This level of prevalence appears higher than the previously reported rates of HCP burnout which are closer to 40%.^3^ Burnout for HCPs working during the COVID-19 pandemic was associated with factors that typically increase the likelihood of HCP burnout. These included feeling pushed beyond training (high workload), making life-or-death prioritizing decisions (high job stress), work impacting the ability to perform household activities (high time pressure), and lack of adequate PPE (limited organizational support).

Burnout among HCPs could be prevented or minimized by actions from healthcare institutions and other governmental and non-governmental stakeholders aimed at potentially modifiable factors. These could include providing additional training and mental health resources, strengthening organizational support for HCPs’ physical and emotional needs, supporting family-related issues (e.g. helping with childcare, transportation, temporary housing, wages), and acquiring PPE. A systematic review showed that both individual- and organizational-level strategies are effective in meaningfully reducing burnout. Some of the most commonly utilized methods focused on mindfulness, stress management and small group discussion.^9^

Recent studies regarding HCPs’ mental health in response to COVID-19 from China, as well as prior studies of other pandemics, have demonstrated that HCPs may experience depression, anxiety, and posttraumatic stress disorder. Shanafelt *et al*. highlighted common sources of anxiety from listening sessions with HCPs that align with our findings, such as access to adequate PPE, unknowingly bringing the infection home, and lack of access to up-to-date information and communication.^10^ HCPs who worked extensively during the SARS pandemic in Beijing later demonstrated posttraumatic stress symptoms (PTSS), and many HCPs in the areas hardest-hit by COVID-19 in China have already started exhibiting similar complaints.^11,12^ To prevent adverse psychological outcomes, mental health support for HCPs is critical.^2,13^Key interventions include access to psychosocial support including web-based resources, emotional support hotlines, psychological first aid, and self-care strategies.

Burnout can impact not only mental health but also can correlate with physical ailments. A systematic review found that burnout was a predictor for conditions including musculoskeletal pain, prolonged fatigue, headaches, gastrointestinal and respiratory issues.^14^ Some factors included in our survey, such as increased workload hours, inadequate PPE or not having updated guidelines, contributed to higher rates of infection among HCP at the beginning of the outbreak in late January.^15^

Burnout was higher in those countries where the COVID-19 pandemic was surging at the time of data collection (e.g. U.S.) compared with those where it was declining (e.g. Italy) or had not reached the peak (e.g. Turkey). The lower reported burnout among HCPs in LMICs may reflect resilience due to more experience working in conditions with high adversity and limited availability of supplies.^16^ Additionally, the greater reported burnout by HCPs in HICs could be attributed to their greater COVID-19 burden. Addressing burnout in all countries is important, but our findings indicate that different strategies should be tailored to the phase of pandemic and the sociocultural and healthcare organizational contexts.

## Limitations

Despite this study’s major strengths, including the breadth of responses from across the globe, there are multiple limitations including a non-validated questionnaire, minimal demographic data collection, and sampling method using social media. Furthermore, drawing comparisons among countries is limited by the differences in cultures and healthcare systems.

## Conclusions

While HCP wage a war against COVID-19, institutions must support these individuals as they face enormous stress that can negatively impact their emotional and physical well-being. Burnout is prevalent at higher than previously reported rates among HCPs working during the COVID-19 pandemic. Reported burnout was significantly associated with, among others, limited access to PPE as well as making life-or-death decisions due to medical supply shortages. Current and future burnout among HCPs could be mitigated by actions from healthcare institutions and other governmental and non-governmental stakeholders aimed at potentially modifiable factors, including providing additional training, organizational support, support for HCPs’ families, PPE, and mental health resources.

## Data Availability

Data will be made available after peer-review publication

## Acknowledgments

The project was supported by the National Center for Advancing Translational Sciences, National Institutes of Health, through Grant UL1TR002003. We acknowledge the support received from Sandra Morales-Mirque, Dr. Craig Niederberger and Dr. Ervin Kocjancic.

## Authors contributions

Drs. Morgantini, Wang, and Weine had full access to all of the data and took responsibility for the integrity of the data and the accuracy of the data analysis.

Study concept and design: All authors.

Literature search: All authors.

Acquisition, analysis, or interpretation of data: All authors.

Drafting of the manuscript: Morgantini, Naha, Vigneswaran, Weine

Critical revision of the manuscript for important intellectual content: Moreira, Abern, Eklund, Weine.

Statistical analysis: Wang, Eklund.

Study supervision: Weine, Crivellaro, Moreira, Abern.

## Conflict of Interest Disclosures

None reported.

## Funding/Support

None reported.

## Role of the Funder/Sponsor

None reported.

## Notes

### Competing Interest Statement

The authors have declared no competing interest.

### Funding Statement

No funding was received for the completion of this study.

